# Epigenetic age acceleration and mortality risk prediction in U.S. adults

**DOI:** 10.1101/2024.08.21.24312373

**Authors:** Angelico Mendy, Tesfaye B. Mersha

## Abstract

**Background:** Epigenetic clocks have emerged as novel measures of biological age and potential predictors of mortality. We aimed to test whether epigenetic age acceleration (EAA) estimated using different epigenetic clocks predict long-term overall, cardiovascular or cancer mortality.

**Methods:** We analyzed data from 2,105 participants to the 1999-2002 National Health and Nutrition Examination Survey aged ≥50 years old who were followed for mortality through 2019. EAAs was calculated from the residuals of Horvath, Hannum, SkinBlood, Pheno, Zhang, Lin, Weidner, Vidal-Bralo and Grim epigenetic clocks regressed on chronological age. Using cox proportional hazards regression, we estimated the hazard ratio (HR) and 95% confidence interval (CI) for the association of EAA (per 5-year) and the DunedinPoAm pace of aging (per 10% increase) with overall, cardiovascular and cancer mortality, adjusting for covariates and white blood cell composition.

**Results:** During a median follow-up of 17.5 years, 998 deaths occurred, including 272 from cardiovascular disease and 209 from cancer. Overall mortality was most significantly predicted by Grim EAA (*P* < 0.0001; HR: 1.50, 95% CI: 1.32-1.71) followed by Hannum (*P* = 0.001; HR: 1.16, 95% CI: 1.07-1.27), Pheno (*P* = 0.001; HR: 1.13, 95% CI: 1.05-1.21), Horvath (*P* = 0.007; HR: 1.13, 95% CI: 1.04-1.22) and Vidal-Bralo (*P* = 0.008; HR: 1.13, 95% CI: 1.03-1.23) EAAs. Grim EAA predicted cardiovascular mortality (*P* < 0.0001; HR: 1.55, 95% CI: 1.29-1.86), whereas Hannum (*P* = 0.006; HR: 1.24, 95% CI: 1.07-1.44), Horvath (*P* = 0.02; HR: 1.18, 95% CI: 1.02-1.35) and Grim (*P* = 0.049; HR: 1.37, 95% CI: 1.00-1.87) EAAs predicted cancer mortality. DunedinPoAm pace of aging was associated with overall (*P* = 0.003; HR: 1.23, 95% CI: 1.08-1.38) and cardiovascular (*P* = 0.04; HR: 1.25, 95% CI: 1.01-1.55) mortality.

**Conclusions:** In a U.S. representative sample, Horvath, Hannum, Pheno, Vidal-Bralo and Grim EAA all predicted overall mortality but only Grim EAA predicted cardiovascular mortality and Horvath, Hannum or Grim EAA predicted cancer mortality. Pace of aging predicted overall and cardiovascular mortality.

## 1. Introduction

Epigenetic modifications or the alterations in gene activity without changes in the DNA sequence potentially transmitted to the organism’s offspring through a process called transgenerational epigenetic inheritance, are a well-known phenomenon associated with aging.^1^ Epigenetic processes include DNA methylation (DNAm), histone modifications and the non-coding RNAs that alter the binding of proteins to DNA and induce or repress gene transcription.^2^ Among epigenetic markers, DNAm is the most stable and quantifiable, it is the selective addition of a methyl group to cytosine within cytosine-phosphate-guanine (CpG) dinucleotide sites to form 5-methylcytosine (5mC).^3^ As early as 1973, Vanyushin et al. reported an inverse relationship between aging and 5-methylcytosine in DNA from rats’ tissues.^4^ These findings were subsequently replicated in various species including humans, leading to the genomic hypomethylation hypothesis of aging, which suggested that reduced 5mC and lower global DNAm occur with aging and may cause a relaxation and abnormal gene expression.^2^ Later research showed that global 5mC does not consistently vary with age; however, DNAm alterations at specific sites of the genome occur with aging.^2^ Such epigenetic alterations are considered as a silent indicator for aging and age associated health risks.^5^

In recent years, epigenetic clocks built from sets of CpGs using mathematical algorithms have emerged as promising measures of biological aging and may improve mortality prediction.^6^ The first generation of epigenetic clocks were developed by Horvarth, Hannum and Weidner to correlate with chronological age.^7-9^ The Horvath clock was developed with DNAm data from various for various human tissues based on 353 CpGs to predict chronological age with 3.6 years mean deviance.^9^ The Hannum and Weidner clocks were based on 71 CpGs and 3 CpGs, respectively and were developed with DNAm data from whole blood.^7,8^ Zhang et al. attempted to improve the performance of these clocks in predicting chronological age by increasing the training sample size, which resulted in the selection of 514 CpGs from whole blood and saliva.^10^

The Vidal-Bralo clock was another first-generation clock, it included 8 whole blood CpGs weighted for their methylation values to predict chronological age.^11^ Second generation clocks such as PhenoAge (513 CpGs) and GrimAge (1030 CpGs) were built with from whole blood DNAm to powerfully predict not only mortality, but also morbidity.^12,13^ The SkinBloodAge clock was developed using 391 CpGs from skin and whole blood to improve DNAm age estimation of previous clocks in fibroblasts and other cell types used in ex-vivo studies.^14^ Lin et al. also produced a clock with 99 whole blood CpGs that correlated with chronological age and cancer.^15^ Third generation clocks such as DunedinPoAm estimate the pace of cellular aging using biomarkers of organ system dysfunction.^16^

Although there are several methods developed to estimate epigenetic clocks in specific conditions, few studies have assessed the ability of epigenetic age acceleration (EAA) to predict all-cause and cause specific mortality in general populations.^17-19^ Therefore, we aimed to determine the association of EAA estimated using epigenetic clocks of first, second and third generation with overall as well as cardiovascular and cancer mortality in a sample representative of the U.S. population aged 50 years or older.

## 2. Methods

### 2.1. Study participants

We used data from the National Health and Nutrition Examination Survey (NHANES) conducted from 1999 to 2002 that included data on epigenetic age. The NHANES is a survey conducted by the National Center for Health Statistics (NCHS) of the Centers for Disease Control and Prevention (CDC) to evaluate the health and nutritional status of U.S. non institutionalized civilian population.^20^ It uses a complex multistage sampling design to derive a sample that is nationally representative.^20^ Written informed consent was obtained from all the study participants and the study design as well as the study protocols were approved by the CDC and NCHS institutional review boards.^20^

Among the 2,532 NHANES 1999-2002 participants who had data on epigenetic age, we excluded participants with missing data on pack-years of cigarette smoking (N = 138), body mass index (BMI) (N = 53), poverty income ratio (BMI) (N = 183). The final sample used for analysis was 2,105 participants.

### 2.2. Epigenetic Clocks

DNA was extracted from whole blood and stored at -80°C until DNA methylation analysis at the Duke University Molecular Physiology Institute. The bisulfite conversion of DNA was performed using a Zymo EZ DNA Methylation kit (Zymo Research, Irvine, California) applying conditions for the Illumina Infinium Methylation assay. DNAm results were produced on the Illumina Infinium MethylationEPIC BeadChi (Illumina, San Diego, California). The data was processed, normalized and used to derive epigenetic biomarkers for the Horvath, Hannum, SkinBlood, Pheno, Zhang, Lin, Weidner, Vidal-Bralo, GrimAge2 clocks. Pace of aging was estimated from the DunedinPoAm. Detailed descriptions of the laboratory procedures, DNAm analysis and quality control are provided at https://wwwn.cdc.gov/nchs/data/nhanes/dnam/NHANES%20DNAm%20Epigenetic%20Biomarkers%20Data%20Documentation.pdf.

### 2.3. Overall, cardiovascular and cancer mortality

The NCHS matched NHANES participants to their National Death Index (NDI) records and also used the death certificates for confirmation. Overall mortality was defined as deaths from any cause, excluding mortality from accident. The specific causes of mortality in our analysis was defined using a standardized list of 113 causes according to the Tenth Revision of the International Classification of Diseases (ICD-10) and included mortality from cardiovascular disease (diseases of the heart) (ICD-10 codes I00-I09, I11, I13, I20-I51) and cancer (ICD-10 codes C00-C97).^21^ The small number of deaths from other causes precluded any further stratification.

### 2.4. Covariates

Baseline information on participants chronological age, sex, race/ethnicity, annual household income, cigarette smoking, physical activity and comorbidities were collected using questionnaires. Using guidelines household income adjusted for family size, year of survey and state, the NCHS estimated PIR.^22^ Participants were asked about moderate to vigorous physical activity in the past 30 days.^23^ BMI was calculated as measured weight in kilograms over height in meter squared.^23^ We defined diabetes as self-reported treatment by oral antidiabetic drug or insulin, fasting plasma glucose ≥126□mg/dl or hemoglobin A_1_C ≥ 6.5%.^23^ We defined hypertension as self-reported use of antihypertension drugs, mean systolic blood pressure ≥ 140□mm Hg or diastolic blood pressure ≥ 90□mm (of up to four measurements on two separate occasions).^23^ Complete blood count measurements were done with the Beckman Coulter method and white blood cell (WBC) differential were performed used VCS technology.^24^

### 2.5. Statistical Analysis

We inspected the intercorrelation between the different epigenetic clocks as well as their correlation with chronological age and pace of aging using Pearson correlations. We performed descriptive analyses to report the distribution of study participants characteristics and we estimated the mean and corresponding standard error (SE) of epigenetic age, epigenetic age acceleration (EAA) and pace of aging. Epigenetic age acceleration (EAA) was calculated for each of the epigenetic clocks by computing the residuals from the regression of epigenetic age on chronological age.^18^ To examined the association of EAA using the different epigenetic clocks and pace of aging with overall and cause-specific mortality, we performed *Cox* proportional hazards regression and estimated the hazard ratio (HR) with corresponding 95% confidence interval (CI). We tested for the proportionality assumption by including interaction terms for each independent variable with follow-up time and none of the interactions were significant, which indicated that no violation was identified.^21^ The models were adjusted for chronological age, PIR, pack-years of cigarette smoking and WBC composition used as continuous variables and gender, race/ethnicity, smoking status, BMI, physical activity, asthma, diabetes, hypertension used as categorical variables. We reported HRs for 5-year increases in EAA and 10% increases in the pace of aging for meaningful interpretation and report of the results. We accounted for the NHANES complex survey design and the sampling weights, so that our results were nationally representative. All analyses were performed in SAS (Version 9.4) and 2-sided *P*-values < 0.05 were considered significant.

## 3. Results

### 3.1. Descriptive results

The study population consisted of 2,105 participants followed for up to 20.7 years (median: 17.5 years [interquartile range: 10.4, 18.7]) during which 998 deaths occurred, including 272 from cardiovascular disease and 209 from cancer. The baseline characteristics of study participants are reported in **Table 1**. In summary, the mean (SE) age of participants was 63.96 (0.34), the majority (54.6%) were female, 79.4% were non-Hispanic White versus 8.3% non-Hispanic Black and 9.1% Mexican-American. About 29.9% had a PIR < 1.85, frequency of current or past smoking was 54.0%, 70.8% were overweight or obese and 53.5% reported moderate or vigorous physical activity in the past 30 days. The prevalence of comorbidities was 5.2% for asthma, 12.1% for asthma and 56.0% for hypertension.

**Table 1:** Baseline Characteristics of study participants (N = 2,105)

| **Characteristics** | **estimate** |
| --- | --- |
| Chronological age (years), mean (SE) | 63.96 (0.34) |
| Epigenetic age (years), mean (SE) |  |
| Horvath | 65.50 (0.30) |
| Hannum | 65.16 (0.29) |
| SkinBlood | 62.16 (0.29) |
| Pheno | 53.67 (0.37) |
| Zhang | 66.11 (0.12) |
| Lin | 55.66 (0.43) |
| Weidner | 53.56 (0.44) |
| Vidal-Bralo | 60.09 (0.25) |
| Grim | 69.98 (0.34) |
| Female sex, % | 54.6 |
| Race/ethnicity, % |  |
| Non-Hispanic White | 79.4 |
| Non-Hispanic Black | 8.3 |
| Mexican-American | 9.1 |
| Hispanic and Other | 3.3 |
| PIR < 1.85, % | 29.9 |
| Smoking, % |  |
| Never | 46.0 |
| Past smoker | 39.1 |
| Current smoker | 14.9 |
| BMI, % |  |
| Underweight (< 18.5 kg/m<sup>2</sup>) | 0.9 |
| Normal (18.5 – 24.9 kg/m<sup>2</sup>) | 28.3 |
| Overweight (25.0 – 29.9 kg/m<sup>2</sup>) | 37.5 |
| Obese (≥ 30.0 kg/m<sup>2</sup>) | 33.3 |
| Physical activity in past 30 days, % | 53.9 |
| Current asthma, % | 5.2 |
| Diabetes, % | 12.1 |
| Hypertension, % | 56.0 |
| EA – chronological age, mean (SE) |  |
| Horvath | 1.54 (0.23) |
| Hannum | 1.21 (0.20) |
| SkinBlood | -1.44 (0.16) |
| Pheno | -10.28 (0.22) |
| Zhang | 2.15 (0.25) |
| Lin | -8.29 (0.26) |
| Weidner | -10.40 (0.39) |
| Vidal-Bralo | -3.87 (0.22) |
| Grim | 6.03 (0.24) |
| DunedinPoAm | 1.10 (0.005) |

Abbreviations: SE = standard error, PIR = poverty income ratio, BMI = body mass index, EAA = epigenetic age acceleration.

On average, epigenetic age was accelerated with the Horvath (mean [SE]: 1.55 [0.23]), Hannum (mean [SE]: 1.20 [0.20]), Zhang (mean [SE]: 2.15 [0.25]), Grim (mean [SE]: 6.03 [0.23]) clocks. However, it decelerated with the SkinBlood (mean [SE]: -1.44 [0.16]), PhenoAge (mean [SE]: -10.28 [0.22]), Lin (mean [SE]: -8.29 [0.26]), Weidner (mean [SE]: -10.40 [0.39]) and Vidal-Bralo (mean [SE]: -3.87 [0.22]) epigenetic clocks **(Table 1**).

### 3.2. Correlation with chronological age and between epigenetic clocks

The strongest correlation between epigenetic clocks and chronological age was observed for Zhang (*r* = 0.90), followed in decreasing order by SkinBlood (*r* = 0.88), Hannum (*r* = 0.88), Horvath (*r* = 0.82), GimAge2 (*r* = 0.82), PhenoAge (*r* = 0.79), Lin, Vidal-Bralo (*r* = 0.66) and Weidner (*r* = 0.60) clocks. Very strong intercorrelations (r ≥ 0.80) were observed between the Horvath, Hannum, SkinBlood, PhenoAge and Zhang clocks. The correlation coefficients of GrimAge2 with the other clocks ranged from 0.65 to 0.80, those of Vidal-Bralo with the other clocks were between 0.65 and 0.47 and those of Weidner with the other clocks were from 0.50 to 0.72 (**Figure 1**).

**Figure 1.**
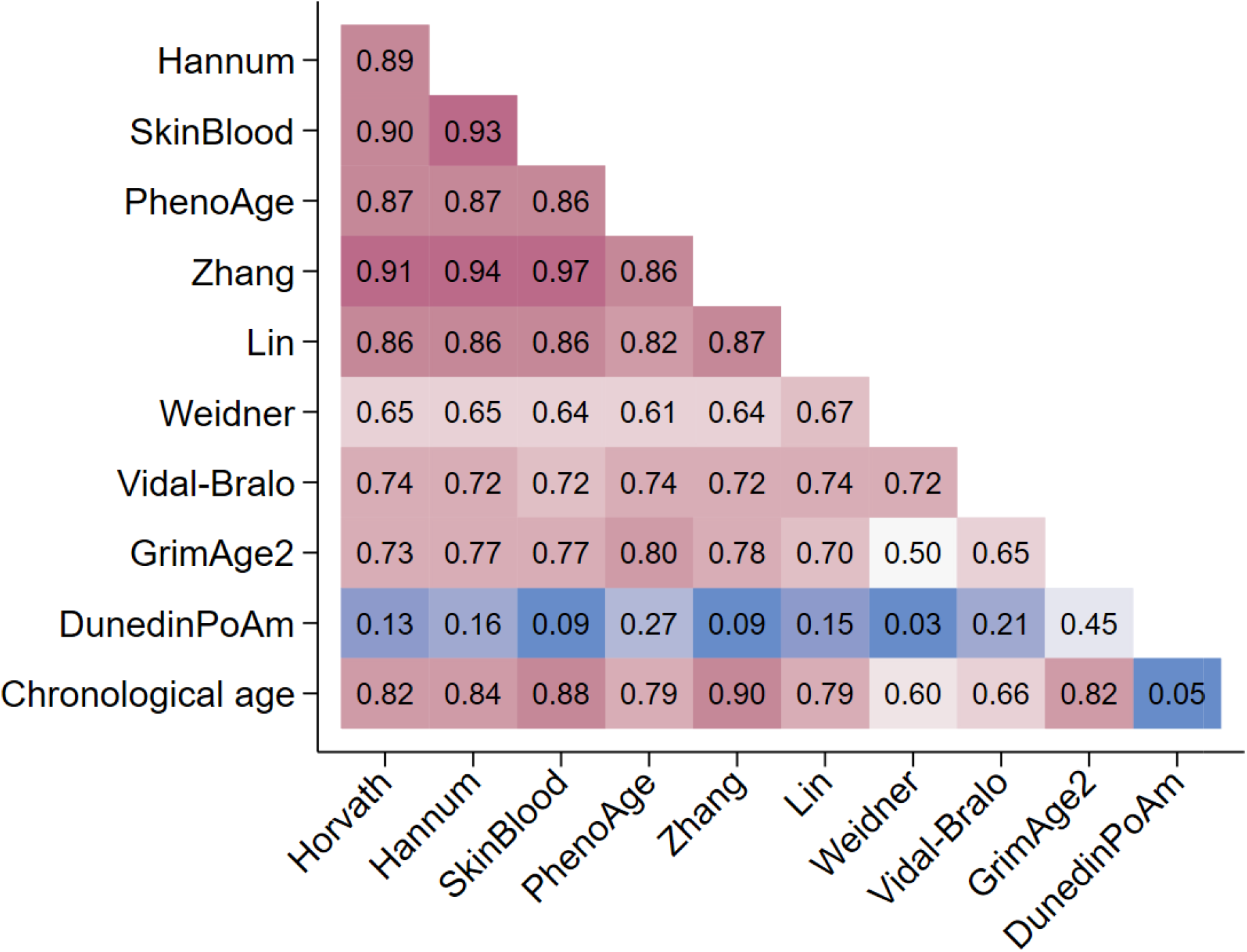
Intercorrelation between epigenetic clocks and correlation with pace of aging and chronological age

### 3.3. EAA and mortality

In Cox proportional hazards regression adjusted for covariates and by order of decreasing significance, higher overall mortality was predicted by increase EAA calculated with the Grim (*P* < 0.0001; HR: 1.50, 95% CI: 1.32, 1.71), Hannum (*P* = 0.001; HR: 1.16, 95% CI: 1.07, 1.27), Pheno (*P* = 0.001; HR: 1.13, 95% CI: 1.05, 1.21), Horvath (*P* = 0.007; HR: 1.13, 95% CI: 1.04-1.22) and Vidal-Bralo (*P* = 0.008; HR: 1.13, 95% CI: 1.03, 1.23) clocks. In cause-specific mortality, only Grim EAA was predictive of cardiovascular mortality (HR: 1.55, 95% CI: 1.29, 1.86; *P* < 0001) whereas Horvath (HR: 1.17, 95% CI: 1.02, 1.35; *P* = 0.03), Hannum (HR: 1.17, 95% CI: 1.02, 1.35; *P* = 0.01) and Grim (HR: 1.37, 95% CI: 1.00, 1.87; *P* = 0.049) EAAs predicted cancer mortality. The DunedinPoAm pace of aging was associated with overall (HR: 1.22, 95% CI: 1.08, 1.38; *P* = 0.003) and cardiovascular mortality (HR: 1.25, 95% CI: 1.01, 1.55; *P* = 0.04) (**Table 2**).

**Table 2.** Epigenetic age acceleration (EAA) and overall, cardiovascular or cancer mortality.

| EAA | Overall mortality (998 deaths) |  | Cardiovascular mortality (272 deaths) |  | Cancer mortality (209 deaths) |  |
| --- | --- | --- | --- | --- | --- | --- |
|  | HR (95% CI) | P-value | HR (95% CI) | P-value | HR (95% CI) | P-value |
| Horvath | **1.13 (1.04, 1.23)** | **0.007** | 1.06 (0.94, 1.19) | 0.33 | **1.18 (1.02, 1.35)** | **0.02** |
| Hannum | **1.16 (1.07, 1.27)** | **0.001** | 1.11 (0.93, 1.33) | 0.23 | **1.24 (1.07, 1.44)** | **0.006** |
| Skin blood | 1.05 (0.96, 1.14) | 0.30 | 1.19 (0.95, 1.50) | 0.12 | 0.97 (0.86, 1.10) | 0.64 |
| Pheno | **1.13 (1.05, 1.21)** | **0.001** | 1.01 (0.91, 1.11) | 0.90 | 1.12 (0.94, 1.33) | 0.20 |
| Zhang | 1.26 (0.97, 1.63) | 0.08 | 1.34 (0.84, 2.14) | 0.21 | 1.10 (0.73, 1.64) | 0.65 |
| Lin | 1.03 (0.98, 1.09) | 0.23 | 1.03 (0.94, 1.12) | 0.53 | 1.03 (0.92, 1.15) | 0.63 |
| Weidner | 1.03 (0.99, 1.07) | 0.10 | 1.02 (0.93, 1.11) | 0.71 | 1.03 (0.93, 1.13) | 0.55 |
| Vidal-Bralo | **1.13 (1.03, 1.23)** | **0.008** | 1.06 (0.91, 1.24) | 0.42 | 1.16 (0.96, 1.41) | 0.12 |
| Grim | **1.50 (1.32, 1.71)** | **< 0.0001** | **1.55 (1.29, 1.86)** | **< 0.0001** | **1.37 (1.00, 1.87)** | **0.049** |
| DunedinPoAm | **1.22 (1.08, 1.38)** | **0.003** | **1.25 (1.01, 1.55)** | **0.04** | 1.07 (0.78, 1.46) | 0.68 |

Abbreviations: HR = hazard ratio, CI = confidence interval, EAA = epigenetic age acceleration.
Hazard ratios calculated using Cox proportional hazards regression. Models adjusted for chronological age, poverty income ratio, pack-years of cigarette smoking and white blood cell composition used as continuous variables and gender, race/ethnicity, smoking status, physical activity, body mass index, asthma, diabetes, hypertension used as categorical variables.
HR reported per 5 year increases in EAA and per 10% increase in DunedinPoAm pace of aging
**Bold** indicates significant association between EAA and mortality outcome

## 4. Discussion

In a sample representation of the U.S. population aged 50 years or older, Horvath, Hannum, Pheno, Vidal-Bralo, and Grim EAAs predicted overall mortality. Cardiovascular mortality was only predicted by Grim EAA and cancer mortality was predicted by Horvath, Hannum and Grimm EAA. Pace of aging was predictive of overall and cardiovascular but not cancer mortality.

This is the first U.S. representative analysis to examine EAA’s association with mortality from specific causes such as cardiovascular disease or cancer and the first to evaluate mortality prediction by EAA developed from the Zhang, Lin, Weidner and Vidal-Bralo epigenetic clocks. A recent analysis of data from 3,581 participants to the Health and Retirement Study (HRS), a U.S. nationally representative study of people aged 50 years or older explored the 4-year all-cause mortality related to Horvarth, Hannum, Pheno and Grim EAAs, but did not include cause-specific deaths.^18^ The study found that all EAAs and the DunedinPACE rate of aging predicted mortality, except the Horvath EAA, after adjusting for sociodemographic characteristics, health behavior and cell types.^18^ In the U.S. Women’s Health Initiative (WHI) study, the association of the Horvath, Hannum, Pheno, and Grim EAAs with healthy longevity defined as survival to age 90 with intact mobility was investigated in 1,813 older women.^17^ However, the study which did not examine cause-specific mortality, found that a standard deviation increase in each of the EAAs was associated with 20% to 40% lower odds of healthy longevity.^17^ In Germany, EAA prediction of cardiovascular, cancer and all-cause mortality was evaluated in 1,864 participants 50 to 75 years old using the Horvath and Hannum clocks, but not second or third generation epigenetic clocks.^19^ In models adjusted for chronological age, sex, batch effects and leukocyte distribution, the Horvath EAA was associated with all-cause, cancer and cardiovascular mortality, whereas the Hannum EAA was only associated with all-cause mortality but not with cancer or cardiovascular mortality.^19^ After further adjustment for smoking, BMI and comorbidities, the Horvath EAA was associated with all-cause and cancer mortality but not with cardiovascular death and the Hannum EAA was not associated with all-cause or cause-specific mortality.^19^ In the Scottish Family Health Study, Grim EAA, but not Horvath, Hannum or Pheno EAAs predicted all-cause mortality in 9,537 followed for 13 years.^25^ Our analysis reported for the first time that EAA developed by Vidal-Bralo in a training set of 390 healthy participants and included 8 whole blood CpGs predicted mortality.^11^ Mortality prediction by this clock has not been previously studied.

In cause-specific mortality, we observed that only Grim EAA predicted cardiovascular mortality. Consistent with this finding, Hillary et al. reported that GrimAge, but not Horvath, Hannum, or Pheno epigenetic clocks predicted incidence of ischemic heart disease in the Scottish Family Health Study.^25^ Similar results were found in African American participants to the Genetic Epidemiology Network of Arteriopathy (GENOA) study where Grim, but not Pheno EAA was associated with cardiovascular disease incidence, independent of traditional risk factors.^26^ The association of Horvath and Hannum EAAs with cardiovascular outcomes has been inconsistently reported in the literature. In the Atherosclerosis Risk in Communities (ARIC) Study, these EAA measures were associated with incident cardiovascular events and cardiovascular mortality among 2,543 African Americans followed for a median of 21 years, when the models were adjusted to chronological age, sex and cell types.^27^ However, the association with cardiovascular mortality became non-significant when the models were additionally adjusted for smoking, physical activity, BMI and covariates such as diabetes or hypertension, all of which were adjusted for in our analysis.^27^ Consistent with our results on cancer mortality related to Horvath, Hannum and Grim EAA, other reported epidemiology studies have reported associations of EAA with incident colorectal cancer, breast cancer in postmenopausal women and lung cancer, especially in smokers.^28-31^ Also similar to our results, pace of aging assessed using DunedinPACE was shown to be associated with overall and cardiovascular mortality and was suggested to mediate the association of life’s essential 8 (diet, physical activity, nicotine exposure, sleep, BMI, blood lipids and glucose and blood pressure) with these outcomes.^32^ The underlying mechanisms for the associations of EAAs with overall and cause-specific mortality are unclear and beyond the scope of the present analysis aimed at identifying the EAA algorithms that best predict these outcomes. EAA may capture long-term exposures associated with adverse health effects and aging as well hormonal, inflammatory and metabolic processes that play a role in disease.^30^ The inconsistent results observed with EAA estimated from various epigenetic clocks may be explained by differences in training data sets, CpGs included in the prediction, the clinical inputs, and statistical methods.^30^

Our analysis had limitations. EAAs were estimated in DNAm extracted from whole blood at a single time point and how they may change over time is unclear. We did not have information on the incidence of cardiovascular disease or cancer and on mortality from specific cancer types. As for any observational studies, residual confounding cannot be completely ruled-out; however, attempts were made to reduce it by adjusting for a wide range of relevant covariates and potential confounders. Nonetheless, this analysis has major strengths. It includes a sample representative of the U.S. older adult population which increases the generalizability of our results. DNA methylation and epigenetic clocks were estimated with rigorous methods and QC using laboratory selected by the CDC.

In conclusion, in a sample representative of the U.S. population aged 50 years or older, Horvath, Hannum, Pheno, Vidal-Bralo and Grim EAAs all predicted overall mortality; however, only Grim EAA predicted cardiovascular mortality, while Horvath, Hannum, and Grim EAA predicted cancer mortality. Pace of aging was predictive of overall and cardiovascular but not cancer mortality.

## Data Availability

All datasets are publicly available and can be accessed through NHANES websites (https://www.cdc.gov/nchs/nhanes/index.htm)

## References

1. Jones MJ, Goodman SJ, Kobor MS. DNA methylation and healthy human aging. Aging cell. 2015;14(6):924–932.

2. Unnikrishnan A, Freeman WM, Jackson J, Wren JD, Porter H, Richardson A. The role of DNA methylation in epigenetics of aging. Pharmacol Ther. 2019;195:172–185.

3. Salameh Y, Bejaoui Y, El Hajj N. DNA methylation biomarkers in aging and age-related diseases. Front Genet. 2020; 11: 171. 2020.

4. Vanyushin BF, Nemirovsky LE, Klimenko VV, Vasiliev VK, Belozersky AN. The 5-methylcytosine in DNA of rats: Tissue and age specificity and the changes induced by hydrocortisone and other agents. Gerontology. 1973;19(3):138–152.

5. Dhar P, Moodithaya SS, Patil P. Epigenetic alterations: The silent indicator for early aging and age-associated health-risks. Aging Med. 2022;5(4):287–293.

6. Protsenko E, Yang R, Nier B, et al. “GrimAge,” an epigenetic predictor of mortality, is accelerated in major depressive disorder. Transl psychiatry. 2021;11(1):193.

7. Hannum G, Guinney J, Zhao L, et al. Genome-wide methylation profiles reveal quantitative views of human aging rates. Mol Cell. 2013;49(2):359–367.

8. Weidner CI, Lin Q, Koch CM, et al. Aging of blood can be tracked by DNA methylation changes at just three CpG sites. Genome Biol. 2014;15:1–12.

9. Horvath S. DNA methylation age of human tissues and cell types. Genome Biol. 2013;14:1– 20.

10. Zhang Q, Vallerga CL, Walker RM, et al. Improved precision of epigenetic clock estimates across tissues and its implication for biological ageing. Genome Med. 2019;11:1–11.

11. Vidal-Bralo L, Lopez-Golan Y, Gonzalez A. Simplified assay for epigenetic age estimation in whole blood of adults. Frontiers in genetics. 2016;7:126.

12. Lu AT, Quach A, Wilson JG, et al. DNA methylation GrimAge strongly predicts lifespan and healthspan. Aging. 2019;11(2):303.

13. Levine ME, Lu AT, Quach A, et al. An epigenetic biomarker of aging for lifespan and healthspan. Aging. 2018;10(4):573.

14. Horvath S, Oshima J, Martin GM, et al. Epigenetic clock for skin and blood cells applied to hutchinson gilford progeria syndrome and ex vivo studies. Aging. 2018;10(7):1758.

15. Lin Q, Weidner CI, Costa IG, et al. DNA methylation levels at individual age-associated CpG sites can be indicative for life expectancy. Aging. 2016;8(2):394.

16. Belsky DW, Caspi A, Arseneault L, et al. Quantification of the pace of biological aging in humans through a blood test, the DunedinPoAm DNA methylation algorithm. Elife. 2020 ;9:e54870.

17. Jain P, Binder AM, Chen B, et al. Analysis of epigenetic age acceleration and healthy longevity among older US women. JAMA Netw Open. 2022;5(7):e2223285.

18. Faul JD, Kim JK, Levine ME, Thyagarajan B, Weir DR, Crimmins EM. Epigenetic-based age acceleration in a representative sample of older americans: Associations with aging-related morbidity and mortality. Proc Natl Acad Sci. 2023;120(9):e2215840120.

19. Perna L, Zhang Y, Mons U, Holleczek B, Saum K, Brenner H. Epigenetic age acceleration predicts cancer, cardiovascular, and all-cause mortality in a German case cohort. Clin Epigenetics. 2016;8:1–7.

20. Centers for Disease Control and Prevention (CDC), National Center for Health Statistics (NCHS), 2018. National Health and Nutrition Examination Survey, Hyattsville, MD. https://www.cdc.gov/nchs/nhanes/about_nhanes.htm, Last accessed date: 16 August 2024.

21. Mendy A, Thorne PS. Long-term cancer and overall mortality associated with drinking water nitrate in the united states. Public Health. 2024;228:82–84.

22. Mendy A, Wilkerson J, Salo PM, Zeldin DC, Thorne PS. Endotoxin clustering with allergens in house dust and asthma outcomes in a US national study. Environ Health. 2020;19(1):1–10.

23. Mendy A. Association of urinary nitrate with lower prevalence of hypertension and stroke and with reduced risk of cardiovascular mortality. Circulation. 2018;137(21):2295–2297.

24. Mendy A, Forno E, Niyonsenga T, Gasana J. Blood biomarkers as predictors of long-term mortality in COPD. Clin Respir J. 2018;12(5):1891–1899.

25. Hillary RF, Stevenson AJ, McCartney DL, et al. Epigenetic measures of ageing predict the prevalence and incidence of leading causes of death and disease burden. Clin Epigenetics. 2020;12:1–12.

26. Ammous F, Zhao W, Ratliff SM, et al. Epigenetic age acceleration is associated with cardiometabolic risk factors and clinical cardiovascular disease risk scores in African Americans. Clin Epigenetics. 2021;13:1–13.

27. Roetker NS, Pankow JS, Bressler J, Morrison AC, Boerwinkle E. Prospective study of epigenetic age acceleration and incidence of cardiovascular disease outcomes in the ARIC study (atherosclerosis risk in communities). Circ Genom Precis Med. 2018;11(3):e001937.

28. Ambatipudi S, Horvath S, Perrier F, et al. DNA methylome analysis identifies accelerated epigenetic ageing associated with postmenopausal breast cancer susceptibility. Eur J Cancer. 2017;75:299–307.

29. Levine ME, Hosgood HD, Chen B, Absher D, Assimes T, Horvath S. DNA methylation age of blood predicts future onset of lung cancer in the women’s health initiative. Aging. 2015;7(9):690.

30. Malyutina S, Chervova O, Maximov V, Nikitenko T, Ryabikov A, Voevoda M. Blood-based epigenetic age acceleration and incident colorectal cancer risk: Findings from a population-based Case–Control study. Int J Mol Sci. 2024;25(9):4850.

31. Dugué P, Bassett JK, Joo JE, et al. DNA methylation-based biological aging and cancer risk and survival: Pooled analysis of seven prospective studies. Int J Cancer. 2018;142(8):1611– 1619.

32. Carbonneau M, Li Y, Prescott B, et al. Epigenetic age mediates the association of life’s essential 8 with cardiovascular disease and mortality. J Am Heart Assoc. 2024:e032743.

